# An artificial-intelligence interpretable tool to predict risk of deep vein thrombosis after endovenous thermal ablation

**DOI:** 10.1101/2024.06.19.24309166

**Authors:** Azadeh Tabari, Yu Ma, Jesus Alfonso, Anthony Gebran, Haytham Kaafarani, Dimitris Bertsimas, Dania Daye

**Affiliations:** Department of Radiology, Massachusetts General Hospital, Boston, Massachusetts, United States of America; Harvard Medical School, Boston, Massachusetts, United States of America; Operations Research Center, Massachusetts Institute of Technology, Cambridge, Massachusetts, United States of America; Trauma, Emergency Surgery, and Surgical Critical Care, Massachusetts General Hospital, Boston, Massachusetts, United States of America

## Abstract

**Introduction:** Endovenous thermal ablation (EVTA) stands as one of the primary treatments for superficial venous insufficiency. Concern exists about the potential for thromboembolic complications following this procedure. Although rare, those complications can be severe, necessitating early identification of patients prone to increased thrombotic risks. This study aims to leverage AI-based algorithms to forecast patients’ likelihood of developing deep vein thrombosis (DVT) within 30 days following EVTA.

**Materials and Methods:** From 2007 to 2017, all patients who underwent EVTA were identified using the American College of Surgeons National Surgical Quality Improvement Program database. We developed and validated 4 machine learning models using demographics, comorbidities, and laboratory values to predict the risk of postoperative deep vein thrombosis: Classification and Regression Trees (CART), Optimal Classification Trees (OCT), Random Forests, and Extreme Gradient Boosting (XGBoost). The models were trained using all the available variables. SHAP analysis was adopted to interpret model outcomes and offer medical insights into feature importance and interactions.

**Results:** A total of 21,549 patients were included (mean age of 54 ± SD years, 67% female). In this cohort, 1.59% developed DVT. The XGBoost model had good discriminative power for predicting DVT risk with AUC of 0.711 in the hold-out test set for all-variable model. Stratification of the test set by age, BMI, preoperative white blood cell and platelet count shows that the model performs equally well across these groups.

**Conclusion:** We developed and validated an interpretable model that enables physicians to predict which patients with superficial venous insufficiency has higher risk of developing deep vein thrombosis within 30 days following endovenous thermal ablation.

## Introduction

There has been rapid growth in the use of endovenous thermal ablation of varicose veins [1]. In 2013, the National Institute for Health and Care Excellence (NICE) recommended endovenous thermal ablation (EVTA) as the preferred treatment option for symptomatic venous insufficiency [2]. This treatment approach induces heat-mediated vessel wall injury, resulting in thrombotic and fibrotic occlusion, which raises concerns about the potential risk of venous thromboembolism (VTE) [3]. Although the complications of deep venous thrombosis (DVT) and pulmonary embolism (PE) are believed to be infrequent, the Society for Vascular Surgery advises patients to undergo early post-procedural duplex scanning to detect potential thrombotic events [4, 5]. Notably, the European Society for Vascular Surgery does not make such a recommendation [6]. The routine use of duplex surveillance has led to the recognition of a new form of localized post-operative DVT which is termed endovenous heat induced thrombosis (EHIT), referring to the extension of thrombus from the ablated superficial vein into the deep vein [7]. EHIT rates reported in the literature vary widely from 0% to 8%, with no consensus on its management [8]. Given the substantial volume of EVTA procedures performed globally and the potential for severe complications, it is imperative for healthcare providers to have a clear understanding of the true incidence of VTE complications [7, 8]. Such insights can aid in individual patient decision-making and facilitate research on VTE prevention strategies [9, 10]. To address these concerns, we have developed machine learning models to predict the 30-day procedure-related risk of DVT in patients undergoing lower extremity endovascular thermal ablation [11]. Leveraging the American College of Surgeons National Surgical Quality Improvement Program (ACS-NSQIP)—a comprehensive, multi-hospital database—we constructed and assessed machine learning models for DVT risk prediction. Each model was analyzed to identify key risk factors, and rigorous testing was conducted to ensure the models’ robustness across diverse demographic groups (race, sex, and age), thus enhancing their applicability in various care settings with differing patient populations.

## Materials and Methods

### Patient selection

A cohort of 21,549 patients who underwent endovenous thermal ablation from January 1, 2007, t o December 31, 2017, across 700 hospitals in the United States was identified using data from th e American College of Surgeons National Surgical Quality Improvement Program (ACS-NSQIP) database [12]. The data were obtained from a national registry, ensuring patient anonymity. The study encompassed procedures performed over a ten-year period. Exclusions from the ACS-NSQIP dataset included minor cases, patients under 18 years old, cases with an ASA score of 6 (brain -death organ donors), trauma cases, transplant cases, cases exceeding the NSQIP contract’s speci fied number, returns to the operating room due to complications of prior procedures, and cases with a prior NSQIP-assessed procedure within thirty days. Data subsets were created using CPT codes to include patients who underwent EVTA for superficial venous insufficiency.

### Definitions and Aim

Patient demographics, laboratory findings and clinical variables including lab values and 30-day postoperative risk of DVT were obtained from the ACS-NSQIP database. The ACS-NSQIP repository also offers a targeted vascular module with pre- and post-operative variables specific to vascular disease and the type of procedure, which was merged with the previously selected cases to provide additional granular details about the procedure performed. A detailed table comprising 2 1,549 observations and 34 variable” was’constructed. This tabular dataset encapsulates diverse features, spanning demographic profiles, clinical particulars, and laboratory outcomes, as delineated in Table 1. The table furnishes an overview of the entire cohort, revealing that the mean (SD) a ge of patients enrolled in model derivation stood at 54.04 (13.7) years. Our primary objective wa s to discern pivotal risk determinants linked with the occurrence of DVT within 30 days post-endovenous thermal ablation (EVTA).

**Table 1.** Patient population demographics and clinical variables.

|  | Training | Testing | Total | *p* |
| --- | --- | --- | --- | --- |
| *N (%)* | 16161 | 5388 | 21549 |  |
| ***Demographics and Race*** |  |  |  |  |
| *Male (%)* | 5320 (32.92) | 1759 (32.6) | 7079 (32.85) | 0.67 |
| *Ethnicity – Hispanic (%)* | 1512 (9.36) | 518 (9.61) | 2030 (9.42) | 0.52 |
| *American Indian or Alaska Native (%)* | 207 (1.28) | 54 (1.0) | 261 (1.2) | 0.07 |
| *Asian (%)* | 560 (3.47) | 176 (3.27) | 736 (3.4) | 0.43 |
| *Black or African American (%)* | 627 (3.88) | 228 (4.23) | 855 (4.0) | 0.18 |
| *White (%)* | 14667 (90.76) | 4890 (90.76) | 19557 (90.8) | 1.00 |
| *Native Hawaiian or Pacific Islander (%)* | 100 (0.62) | 40 (0.74) | 140 (0.6) | 0.25 |
| ***Clinical Comorbidities and Presenting Symptomatology*** |  |  |  |  |
| *Age (mean (SD))* | 54.04 (13.81) | 54.03 (13.73) | 54.04 (13.79) | 0.98 |
| *ASA Class 1 – No disturb (%)* | 2932 (18.14) | 973 (18.06) | 3905 (18.12) | 0.68 |
| *ASA Class 2 – mild disturb (%)* | 9574 (59.24) | 3210 (59.58) | 12784 (59.33) | 0.68 |
| *ASA Class 3 – severe disturb (%)* | 3485 (21.56) | 1144 (21.23) | 4629 (21.48) | 0.68 |
| *ASA Class 4 – life threat (%)* | 166 (1.03) | 61 (1.13) | 227 (1.05) | 0.68 |
| *ASA Class 5 – moribund (%)* | 4 (0.02) | 0 (0.00) | 4 (0.02) | 0.68 |
| *Smoker (%)* | 1924 (11.91) | 677 (12.56) | 2601 (12.07) | 0.14 |
| *COPD (%)* | 315 (1.95) | 102 (1.89) | 417 (1.94) | 0.77 |
| *Ascites (%)* | 4 (0.02) | 1 (0.02) | 5 (0.02) | 0.77 |
| *CHF (%)* | 32 (0.20) | 14 (0.26) | 46 (0.21) | 0.31 |
| *Hypertension (%)* | 5522 (34.17) | 1816 (33.70) | 7338 (34.05) | 0.47 |
| *Renal failure (%)* | 9 (0.06) | 1 (0.02) | 10 (0.05) | 0.25 |
| *Bleeding disorder (%)* | 494 (3.06) | 159 (2.95) | 653 (3.03) | 0.65 |
| *Transfusion requirement (%)* | 5 (0.03) | 2 (0.04) | 7 (0.03) | 0.80 |
| *BMI (mean(SD))* | 30.47 (7.66) | 30.38 (7.58) | 30.45 (7.64) | 0.44 |
| ***Pre-Op Labs (mean(SD))*** |  |  |  |  |
| *Sodium* | 139.57 (2.07) | 139.59 (2.10) | 139.58 (2.07) | 0.58 |
| *BUN* | 15.75 (5.75) | 15.73 (5.88) | 15.75 (5.78) | 0.78 |
| *Creatinine* | 0.87 (0.50) | 0.87 (0.47) | 0.87 (0.49) | 0.75 |
| *WBC* | 6.63 (1.72) | 6.64 (1.83) | 6.63 (1.75) | 0.56 |
| *Hematocrit* | 40.58 (3.41) | 40.51(3.46) | 40.56 (3.43) | 0.20 |
| *Platelet* | 239.94 (55.07) | 240.50 (54.86) | 240.08 (55.01) | 0.51 |

Four machine learning models were evaluated for their ability to predict 30-day risk of DVT after EVTA as the outcome: Classification and Regression Trees (CART), Optimal Classification Trees (OCT), Random Forests, and Extreme Gradient Boosting (XGBoost).

### Missing Data Imputation

Of the 34 features in the data, 17 had missing values. It’s been noted that missing values in datas ets are often not randomly distributed [13], which could lead to biased results if patients with incomplete data are removed. To avoid this, the missing values were filled in using Optimal Imputation, a method that has been shown to perform better than other state-of-the-art optimization techniques [14].

### Training/Testing Data Splits

The dataset was then divided into two sections – training data, which comprised of 16,161 cases, and independent testing data, consisting of 5,388 cases. This division follows the standard machine learning practice of a 75/25 split, where the majority of the data is allocated for training purposes, while a portion is reserved for evaluation.

### Grid-search Model Training

We use the training data to train potential model candidates including CART, random forest, Opt imal Classification Trees and gradient boosted trees with different hyperparameters. These model s are selected since they are all tree-based methods that will allow more interpretability access fo r clinical use. We also aim to compare all these models to select the most state-of-the-art model f or this particular task. Class imbalance, where one class has significantly more samples than the other, can negatively impact model performance and lead to biased results where the majority class is overrepresented. To address this, we employed sample weighting for each model, whichh ar tificially increases the weight of the minority class. This helps ensure a more balanced representa tion of both classes in the model. Finally, we test all possible hyperparameter combinations for e ach model by conducting a grid search and selecting the combination that achieved the highest A UC. The model was finally then evaluated using Area under the Receiver operator curve (AUC R OC), sensitivity, and specificity.

The parameters that were tuned include: for C–RT - maximum tree depth and minimum number of samples per leaf; for Random For–st - maximum tree depth, minimum number of samples per leaf, number of trees in the forest, and cost complexity parameter; for Optimal Classification Tr– es - maximum tree depth and minimum number of samples per leaf; and for Gradient Boosted Tr –es - number of tree estimators, step size shrinkage, and minimum loss reduction required to mak e a further partition on a leaf node of the tree.

### SHAP Interpretability Analysis

The selected best model was interpreted using Shapley Additive exPlanations (SHAP). SHAP is a game-theoretic approach that explains the output of a machine learning model by quantifying t he individual contributions of each feature to the final prediction, both for individual samples an d the overall population. This method also allows for the examination of linear and nonlinear interactions between features [15].

### Software Requirements

These models were developed and evaluated using Python version 3.8.13 (packages: interpretableai, numpy, pandas, scikit-learn, xgboost, shap, math). The full procedure is illustrated in Fig 1. This study was approved by the Mass General Brigham Institutional Review Board. The data were analyzed anonymously, and consent was waived for this study.

Fig 1. Criteria for patient inclusion.

### Statistical Analysis

Data processing and analysis were performed using Python 3.8.13. Summary statistics were presented as total counts and frequencies for categorical variables and as mean and standard deviation values for continuous variables. The differences of data distribution between data sets for both categorical and continuous variables were assessed by Chi-square test and t-test, respectively.

### Ethics Statement

Ethical standards were upheld throughout the study and approved by the institutional review board of Massachusetts General Hospital. A waiver of consent was obtained from the institutional review board committee due to the use of de-identified patient data and no identifiable medical risk, relying solely on pre-existing information in patient medical records.

## Results

### Cohort

Our study comprised a total of 21,549 patients. Table 1 provides characteristics of both the training and testing cohorts, as well as the entire study population. On average, patients included in the model development had a mean (SD) age of 54.0 (13.8) years and a mean BMI of 30.45 (7.64) kg/m². The majority of individuals in the cohort were female, accounting for 67.2% of the total, while 22.5% were categorized as ASA class ≥ 3. Additionally, 12% of the cohort reported a history of smoking.

### Model Performance

The top-performing machine learning model for predicting the 30-day risk of deep vein thrombosis (DVT) following endovenous thermal ablation (EVTA) was XGBoost. This model demonstrated an AUC-ROC of 0.711, with an accuracy of 0.985, sensitivity of 1.00, and specificity of 0.047 when evaluated on the testing set. The results of all models on the training and testing sets are shown in Table 2.

**Table 2.** Comparison of model performance and metrics of the four models.

| Model | AUC in Train Set | AUC in Test Set | Accuracy | Sensitivity | Specificity |
| --- | --- | --- | --- | --- | --- |
| OCT | 0.759 | 0.580 | 0.616 | 0.547 | 0.617 |
| CART | 0.637 | 0.558 | 0.680 | 0.419 | 0.684 |
| Random Forest | 0.789 | 0.670 | 0.767 | 0.442 | 0.773 |
| Gradient Boosted Trees (XG Boost) | 1 | 0.711 | 0.985 | 1.000 | 0.047 |

The AUC-ROC curve is pictured in Fig 2. This model incorporated thirty-four features, including demographic variables, physiological high-risk factors, and various laboratory parameters such as hematocrit, blood urea nitrogen (BUN), creatinine, white blood cell count, platelet count, elective surgery status, renal comorbidities, dyspnea, and presence of open wound or wound infection, among others (detailed in Table 1). The random forest model, an ensemble machine learning approach comprising numerous individual decision trees, operates independently to generate predictions, with the collective output of all trees averaged for final prediction. Figure 3 presents the SHAP summary plot, highlighting features deemed most influential in the model. Notably, BMI, Preoperative Platelet count, and age emerge as prominent contributors. Additionally, the coloration of features indicates the direction of their impact; for example, higher values (red) in Preoperative Platelet count signify an elevated SHAP value, corresponding to an increased risk of deep vein thrombosis.

Fig 2. Area under the receiver operating curve for XGBoost model.

Fig 3. Feature importance plot for all-variable model, where the red indicates a higher feature value, and blue indicates a smaller feature value. The features are ranked by their contribution importance to the model, and thus BMI is considered the most important feature. The rest of the 25 features not demonstrated have their aggregated importance summarized as the last point.

In addition, Fig 4 demonstrates SHAP dependence plot of BMI, preoperative platelet count, age and preoperative WBC count, top 4 most important features from the gradient boosted trees model versus its SHAP value. For example, for the age dependence plot (plot 3A), as age increases above a threshold of 46, patients become more likely to develop DVT, although this likelihood starts decreasing again at around 60 years old. Positive SHAP values indicate that this feature value contributes to the positive prediction of developing DVT, while negative SHAP indicate a contribution to predicting no DVT event.

Fig 4: SHAP dependence plots of the top four importance features.

### An Example of the Power of Personalized use of the Predictive Model

One of the main advantages of our proposed approach is its ability to demonstrate the effects of predictive features on each individual outcome. Using SHAP, we can see how each feature contributes to the prediction of outcome for each instance in the data. This contribution will include both the direction and the magnitude of the variable effect given the feature values for that particular sample. In this section, we highlight the significant benefit of employing the proposed personalized usage of the predictive model based on an example (Fig 5): a 67-year-old woman, self-identified as White, was admitted with an ASA class 2, had a BMI of 25 and showed no abnormal clinical variables. For this patient the SHAP plots of XGBoost model performance using all variables exhibits the log of the population level probability of having DVT (E[f(X)]), and each bar represents the contribution of each feature to either increase or decrease the probability of having DVT. We observe that all features represented as most important for this patient also correlates to the population-level most important features. The feature contribution direction also corresponds well to the previous SHAP interaction plots, where having age of 67 pushed the probability of DVT negative by log (-0.21) and having BMI of 25.25 similarly pushed the probability by log (-0.91). Meaning when the patient is high aged and high BMI, their risk of DVT in fact decreases.

Fig 5: Visualization with SHAP plots of XGBoost model performance using all variables in an example patient.

### Difference between the two models

We observe that the order of top contributing features that are selected in the two examples of the model based on the whole population and on a single patient are different. The reasoning for this is although certain features aggregately are more important than others, when down to individual patients, depending on their other various characteristics, the effect of influence of those features are unique to that patient. From a technical point of view, recall that each individual follows potentially different decision paths of the decision trees among XGBoost and random forest models. This means that different individuals’ final predicted outcome may depend on different features. Although certain features are consistently considered importantly aggregately across the entire population, others become more specific and dependent on individual patients. For example, patient A’s decision path only follows Preop Platelet, and thus SHAP indicates these two are most significant for this patient. However, aggregately, other patients strongly rely on BMI and Preop Platelet, as well as others, then SHAP would indicate a different ranking for the full cohort.

## Discussion

In this study, we have developed four machine learning models aimed at predicting the risk of deep vein thrombosis occurrence within 30 days following EVTA. The random forest and XGBoost models were shown to perform well (AUC: 0.67 and 0.71, respectively). We interpreted the models using SHAP in order to gain an understanding of the importance and direction of influence of each feature. Among patients undergoing lower extremity endovascular thermal ablation, the most important predictor of DVT was age, BMI, hypertension and preoperative lab values. For each of the top four features, we split the cohort into subgroups and performed SHAP analysis, to identify differences between plots. We compared the performance of several different machine learning models on the task of predicting 30-day procedure-related risk of DVT development: OCT, CART, Random Forests, and XGBoost. The CART and OCT performed similarly on the AUC metric, but the specificity and accuracy of CART model were higher at 0.68. The specificity of XG Boost was the lowest at 0.047, which indicates that this model generated a large number of false positives. Conversely, the Random Forest achieved high specificity but low sensitivity, demonstrating an inability to perform the key task of identifying patients at risk of DVT development.

For 30-day risk prediction, the XG Boost model achieved the highest AUC at 0.711, with good sensitivity and accuracy compared to the OCT, CART and Random Forest, which have low sensitivity at 0.54, 0.41 and 0.44, respectively. These findings suggest that XGBoost, as a tree-based method, could be the most appropriate choice for predicting the risk of DVT. This preference may stem from the method’s capability to account for nonlinear relationships among variables present in the ACS-NSQIP database.

While prognostic tools for venous thromboembolism exist for patients with various underlying conditions [16, 17], to the best of our knowledge, this study represents the first attempt to utilize an AI-based tool for predicting the risk of developing DVT following EVTA. In a prior study focusing on the early diagnosis of DVT in patients undergoing hip arthroplasty, XGBoost performed as the top-performing model, achieving an impressive AUC of 0.982 [18].

Conventional approaches to identifying risk factors typically involve constructing risk models using univariate or multivariate regression techniques. However, these methods often fail to account for interactions and nonlinear relationships among variables [18]. In contrast, ML models have demonstrated superior capability in capturing complex relationships among variables, thereby uncovering potential nonlinear associations and interactions. Ensuring model interpretability is crucial in medical decision-making applications of machine learning, as transparency remains a significant practical concern in the integration of AI into clinical workflows. Methods for model interpretability also empower clinicians to uncover novel clinical insights from machine learning models. One such technique is the SHAP algorithm, utilized for ML interpretation. Grounded in the Shapley value concept from game theory, this algorithm computes the contribution of each feature to the prediction. This allows us to understand the impact of each feature on the model and further explain the model’s prediction results. In this study, we leverage SHAP explanatory methods to understand both the importance of each feature across all predictions as well as the direction of influence of each feature for individual predictions following model development. The key variables we have identified align closely with established knowledge regarding the factors contributing to post-EVTA risks of DVT development, underscoring the utility of our model. For example, physiologic high-risk factors including body mass index and age, and preoperative WBC count and platelet value are the top 4 most important predictors for DVT development. The well-documented association between DVT and factors such as age and weight further validates our model’s recognition of physiological high-risk factors as pivotal predictors [19–22].

Other important predictors for DVT, preop platelet and WBC count, also demonstrate the clinical relevance of the model. Studies in the 1970s using radiolabeled leukocytes have shown uptake of white blood cells into venous thrombi, while accumulation of polymorphonuclear neutrophils (PMNs) on the abluminal side of the endothelium following occlusion of veins, first led to the speculation that ‘white-cell’ induced endothelial damage in contributing to venous thrombosis in humans [23]. The exposure of the collagen-rich wall is purported to trigger platelet aggregation and subsequent leukocyte sequestration, creating a nidus for thrombus propagation [24]. While the role of WBCs in the natural progression of venous thrombus is multifaceted, recent findings indicate that recruited PMNs may instigate thrombosis by generating neutrophil extracellular traps [23, 25].

Our model emphasizes the significance of these variables and others, providing interpretable machine learning insights into the 30-day procedure-related risk of DVT development. We find the results obtained from our XGBoost model to be promising, particularly given its consideration of various patient characteristics. However, we acknowledge the possibility of overfitting due to imbalanced data distribution and advocate for precautionary measures to address this concern. Nonetheless, our study has several limitations. The reliance on the ACS-NSQIP database, which is predominantly utilized in large teaching hospitals with greater resources for data collection, may introduce biases and limit generalizability [25]. Therefore, the data available in the ACS-NSQIP database may not fully represent the spectrum of surgical cases across the United States. Moreover, the database solely captures 30-day outcomes, precluding the examination of longer-term DVT risk. ACS-NSQIP also lacks certain clinically relevant variables such as the intervened vessel, operator expertise, and procedural variations. Another limitation stems from the CPT code filtering process, assuming uniformity across all procedures. Furthermore, the observed disparities in model performance between training and testing sets suggest a degree of overfitting to the training data. This phenomenon could be attributed to the imputation of synthetic data from a limited pool of known observations [14, 26]. However, our models demonstrate robust performance on an independent test set, suggesting that this concern may be somewhat mitigated. It’s important to note that the feature selection and interpretation steps utilizing SHAP only establish correlations between the selected variables and the investigated outcomes, and cannot infer causal relationships. In addition, technical limitations include a class imbalance issue, with only 1.59% of the patient population experiencing a positive outcome, potentially biasing the algorithm training process towards negative samples. To address this, we implement balancing weighting techniques in various stages of the modeling process to ensure parity between positive and negative classes.

## Conclusion

We have developed and validated an XGBoost interpretable model that enables physicians to predict which patients with superficial venous insufficiency has higher risk of developing deep vein thrombosis within 30 days following endovenous thermal ablation. This model may help us personalize the medical decisions to minimize the risk of DVT development in these patients.

## Author Contributions

Y.M. led the efforts of data processing, planning, and performing experiments, model development and implementation, writing code, analyzing, and validating results, and wrote the manuscript. A.T. validated results and wrote the manuscript. J.A. contributed processed data, wrote codes, and performed experiments. A.G. collected data, validated results and edited manuscript. H.K. contributed to experimental design, validated results and edited manuscript. D.D supervised and led the medical team, contributed to experimental design and model evaluation, and edited the manuscript. D.B. directed the overall project, from concept and research to implementation, and edited the manuscript.

## Data Availability

All data produced are available online at the American College of Surgeons National Surgical Quality Improvement Program database

https://www.facs.org/quality-programs/data-and-registries/acs-nsqip/

